# Vestibular agnosia is linked to worse balance recovery in traumatic brain injury: a longitudinal behavioural and neuro-imaging study

**DOI:** 10.1101/2024.02.12.24302715

**Authors:** Zaeem Hadi, Mohammad Mahmud, Elena Calzolari, Mariya Chepisheva, Karl A Zimmerman, Vassilios Tahtis, Rebecca M Smith, Heiko M Rust, David J Sharp, Barry M Seemungal

**Affiliations:** Centre for Vestibular Neurology (CVeN), Department of Brain Sciences, Charing Cross Hospital, Imperial College London, London, W6 8RF, UK; Department of Brain Sciences, Hammersmith Hospital, Imperial College London, London, W12 0NN, UK; UK Dementia Research Institute, Care Research & Technology Centre, Imperial College London, London, UK; King’s College Hospital NHS Foundation Trust, London, SE5 9RS, UK; Department of Neurology, University Hospital Basel.

**Keywords:** vestibular recovery, traumatic brain injury, self-motion perception, postural balance, resting-state functional connectivity, diffusion tensor imaging

## Abstract

**Background and Objectives:** Moderate-to-severe traumatic brain injury patients commonly manifest vestibular dysfunction with imbalance and dizziness. Importantly, falls in these patients are linked to long-term unemployment and increased mortality. There are however no objective acute-prospective longitudinal data of vestibular outcomes nor of the mechanisms predicting vestibular recovery. We previously showed that in acute traumatic brain injury, acute imbalance was linked to impaired vestibular perception of self-motion (i.e. vestibular agnosia) via right inferior longitudinal fasciculus disruption. This report importantly could not inform upon the predictors nor neural substrates of vestibular recovery, questions which we focus upon in this first acute-longitudinal study assessing objective vestibular recovery in traumatic brain injury, with detailed clinical, laboratory and neuroimaging measures.

**Methods:** We screened 918 hospitalized traumatic brain injury patients, recruiting 39 acutely (18 – 65 age) with preserved peripheral vestibular function confirmed via vestibular ocular reflex testing (patients 2.52°/s versus controls 1.78°/s; *P* > 0.05) of which 34 patients (17 with vestibular agnosia) completed the follow-up testing. Common inner ear and migraine diagnoses were treated to resolution pre-testing. Vestibular perceptual thresholds were assessed via whole-body rotations in the dark on a computerized rotating chair. Using k-means clustering, vestibular perceptual thresholds of healthy controls and patients were compared resulting in 1.99 °/s/s or above being classified as vestibular agnosia. Balance was assessed via force platform.

**Results:** The change in vestibular perceptual thresholds and sway from acute to follow-up was linked (*P* < 0.05) and a logistic model indicated (*P* < 0.05) that those who had acute vestibular agnosia made worse recovery of vestibular mediated function (balance and vestibular perception) as compared to non-vestibular agnosia patients. Moreover, subjective symptoms of imbalance and dizziness (via ‘Dizziness Handicap Inventory’) were not linked to objective vestibular recovery of balance and perception (*P* > 0.05). Neuroimaging findings indicated that the linked vestibular recovery (from vestibular agnosia and imbalance) was mediated by bihemispheric fronto-posterior circuits particularly frontal poles and splenium of corpus callosum.

**Conclusion:** Acute vestibular agnosia predicts worse recovery of imbalance and linked recovery of vestibular agnosia and is mediated by partially overlapping, bi-hemispheric circuits. Since vestibular dysfunction may occur without vertigo complaint (from vestibular agnosia), clinical and research assessment of vestibular recovery should assess objective vestibular measures in addition to dizziness symptoms, which poorly track central vestibular recovery. Finally, our cohort were young and without medical co-morbidities, hence understanding the additional impacts of ageing and multimorbidity is required in vestibular recovery in traumatic brain injury cohorts.

## Introduction

Traumatic brain injury (TBI) is the leading cause of chronic disability and death in young adults,^1^ and the cause of death in half of over-65-year-olds dying post-unintentional fall.^2^ Vestibular dysfunction with dizziness and/or imbalance, affects c. 80% of acute TBI patients,^3^ and c. 50% of TBI survivors at 5 years.^4^ Typical vestibular disorders include vestibular agnosia (‘VA’) i.e., attenuated sensation of self-motion perception, imbalance and dizziness, to inner ear diagnoses such as benign paroxysmal positional vertigo (‘BPPV’), which could result in mild to intense episodes of dizziness. Persisting vestibular dysfunction in TBI survivors is linked to falls,^5^ potentially explaining increased long-term mortality. Hence, understanding the mechanisms of recovery from imbalance in TBI, may enable identification of patients at risk of falls, and developing and monitoring targeted therapy for imbalance and falls in TBI.

Prospective data assessing objective vestibular outcomes in TBI are however sparse,^6^ an important omission since damage to perceptual mechanisms (e.g. VA), uncouples symptoms from signs, and results in a lack of correlation between subjective symptoms and objective deficit (supplementary video – shows a patient reporting no sensation of dizziness in presence of a strong nystagmus in response to caloric irrigation).^3,7,8^ This uncoupling of symptoms from signs was also previously noticed in TBI patients c. 90 years ago^9^ by Glaser who noted in a series of 66 traumatic brain injury patients, “*Exceedingly difficult to understand, however, is the absence of true vertigo in head injuries, in spite of the presence of peripheral and central vestibular damage*”. This conundrum remained unexplained until recent work in elderly individuals^10–15^ with cortical white-matter lesions, postural imbalance, and attenuated vestibular perception (i.e., VA), even during strong vestibular stimulation via caloric irrigation.^12,13^

Our recent work, followed by above reports in the elderly, showed the first evidence of vestibular agnosia in TBI, which explains the lack of concordance in symptoms and vestibular function observed previously.^3,8,9^ Despite higher prevalence of BPPV, the TBI patients,^7,16^ same as elderly individuals,^11,17,18^ have fewer dizziness complaints due to VA.^11–13,17^ Hence, we predict that using subjective report of symptoms and not accounting for VA, will inaccurately track vestibular dysfunction in TBI. For example, BPPV prevalence in similar in-patient rehabilitation TBI cohorts found prevalence of 7%^19^ based upon screening via symptoms (dizziness) versus a 58% BPPV prevalence^16^ based upon a systematic examination irrespective of symptoms. Thus, subjective symptom scores underdiagnose vestibular conditions in TBI cohorts.

In contrast to subjective vestibular symptoms which did not predict objective deficits in acute TBI,^7^ laboratory measures of VA^10^ were linked to worse laboratory measured balance. Acutely, using neuroimaging we showed that this VA–imbalance overlap was mediated by disruption to the right inferior longitudinal fasciculus. However, key questions unresolved by our first report included the temporal pattern of recovery of dizziness symptoms, the clinical significance of VA recovery for balance recovery, and the neuroanatomical correlates of vestibular recovery which may differ from that mediating the overlap in acute VA and imbalance (i.e. the right inferior longitudinal fasciculus – ‘ILF’).^7^

Our focus on vestibular recovery was motivated by clinical need since balance recovery is linked to return-to-work rates in TBI,^5,20^ and, via imbalance-related falls,^2^ to excess mortality in community-dwelling TBI survivors.^21^ Hence, using a priori hypotheses linking vestibular agnosia with imbalance, we conducted the first acute, prospective, longitudinal study assessing objective vestibular dysfunction in TBI patients. As such, this report provides new longitudinal data relevant to recovery of vestibular mediated balance and perception, in a cohort we previously reported only acute data.^7,22,23^

## Methods

### Cohort recruitment and testing

Approval was obtained by the local Research Ethics Committee (17/LO/0434). The principles of the Declaration of Helsinki were adhered to. Recruitment details, previously reported,^7^ are briefly mentioned here.

Following informed consent, testing occurred during the patients’ admission or immediately after discharge, 3months and 6months. As per our approved protocol, patients without capacity at acute testing were recruited via a consultee and informed patient consent was obtained subsequently. Prior to laboratory testing, any BPPV and/or residual migraine-phenotype headaches were treated to resolution via repositioning manoeuvre (BPPV) and medication (migraine: 3–5 days of Naproxen and Prochlorperazine).^7^

**Inclusions:** (i) in-patient on a major trauma ward with blunt head injury; (ii) age 18–65; (iii) preserved peripheral vestibular function (nb: and laboratory confirmed in all tested patients since peripheral vestibular hypofunction affects c. 15% of acute TBI).^3^

**Exclusions:** (i) active pre-morbid medical, neurological, or psychiatric conditions; (ii) musculoskeletal condition impairing balance; (iii) substance abuse; (iv) pregnancy; and (v) inability to obtain consent or assent. Age and sex matched healthy controls were tested once.

### Procedure

Comprehensive testing (0, 3, 6-months) included evaluating peripheral and reflex vestibular function (vHIT), vestibular perceptual testing, posturography, and neuroimaging (MRI at 0, 6months). Bithermal caloric irrigation or electronystagmography (ENG) with rotational chair testing was performed if a video head impulse test was not possible due to neck pain.

### Assessment of peripheral and reflex vestibular function

For VOR thresholds, participants sat on a computer-controlled vibrationless rotating chair in dark, which accelerated by 0.3 deg/s^2^ from stop, every 3s for 33s, and then returned to stop (detailed algorithm previously described).^7^ The chair velocity which elicited the first clearly observable nystagmus was recorded resulting in VOR thresholds of patients: 2.52°/s versus controls: 1.78°/s indicating normal reflex thresholds (t = 1.742, *P* > 0.05). Details from vHIT, caloric, and ENG testing are reported in supplementary table (Table S1) and are also previously reported,^7^ all of which indicated that patients had normal peripheral vestibular function.

### Vestibular perceptual thresholds

Vestibular perceptual thresholds (VPTs) of perceived self-motion during passive, yaw-plane, whole-body rotations in the dark were obtained via a previously described iterative staircase algorithm.^7,10^ Briefly, participants were passively rotated (left or right) in the dark and with white noise sound masking, while sitting on a motorised and vibrationless computer-controlled rotating chair (Contraves, USA). The rotation kinematics were within the optimal range for semi-circular canal transduction of head angular motion, with constant acceleration rotations of duration 5s (i.e., frequency 0.2Hz).^24^ Subjects were instructed to press a right or left button as soon as they perceived the rotation in the respective direction. If the button was pressed indicating the correct direction and within the 5s of the rotation start, the algorithm registered the response as ‘correct’ and the next rotation in that direction was lowered in acceleration. If there was no button press response or an incorrect response (indicating the wrong direction) before 5s, then (a) the chair started decelerating to stop (b) the response was registered as ‘wrong’. Thus, the next rotation trial in that direction was increased in constant acceleration. The algorithm could backtrack to account for mistakes or chance correct responses as previously described.^7,10^ Importantly, the lights were turned on after each rotation to ‘dump’ unwanted post-rotatory vestibular effects. The apparatus and an example of the recorded signals are shown in (Fig 1A-1B).

**Figure 1.**
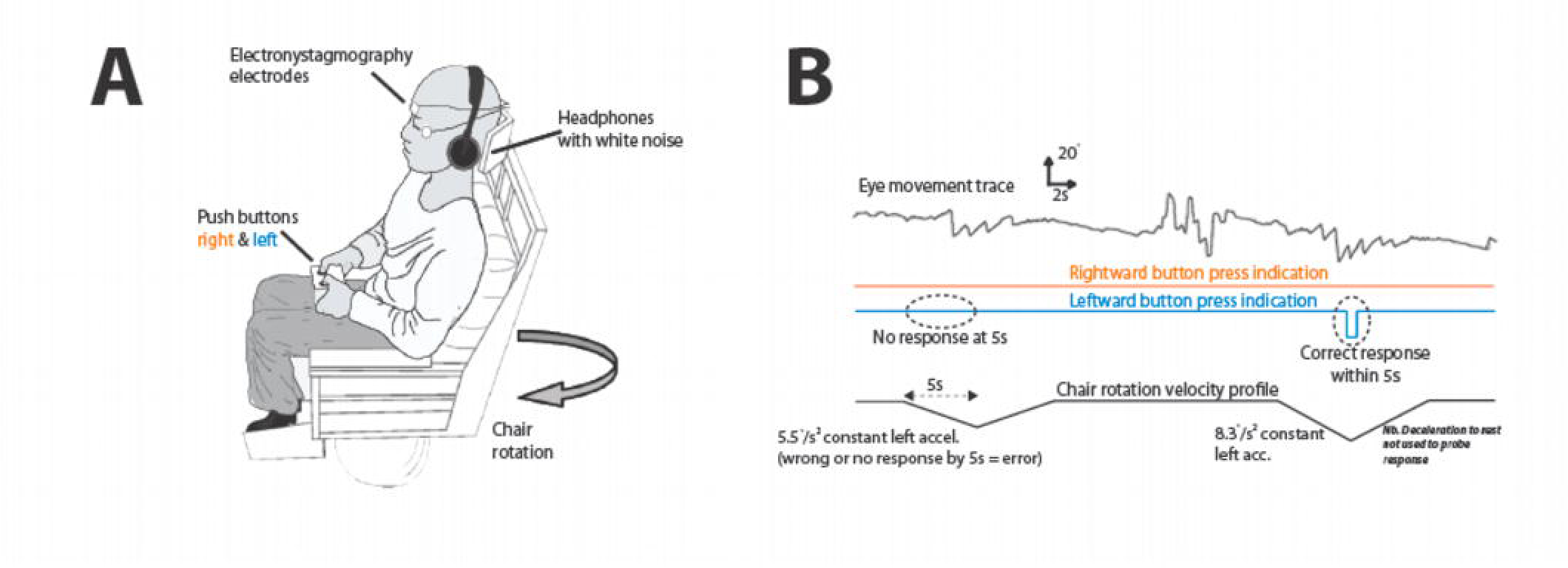
Vestibular agnosia measurement. **(A)** Patients were seated on a rotating chair in dark and rotated in yaw plane and were instructed to press right or left to indicate rightward or leftward rotation. Background noise was masked using white-noise whereas eye-movements were also recorded. **(B)** Traces indicating eye-movements, button-press by participant, and velocity profile of computer-controlled chair rotation. *(see supplementary video for demonstration of vestibular agnosia during caloric irrigation test)*

Acutely, our patient cohort were categorized into vestibular agnosia (VA+) or no vestibular agnosia (VA-) by comparison with healthy controls’ vestibular perceptual thresholds (mean: 0.76 °/s^2^; SD: 0.42) using k-means clustering, which resulted in 1.99 °/s^2^ or above acceleration threshold on either side (right or left sided rotations) being classified as vestibular agnosia.^22^

### Posturography

Postural sway was measured via a force platform. We analysed sway during the condition “soft surface-eyes closed” (SC) when balance is heavily reliant upon vestibular cues since vision and proprioception is absent or degraded. We showed previously that condition SC best discriminated TBI patients from controls.^7^ The root mean square (RMS) of sway for SC was used for all balance analyses in this report. All postural sway data was processed using custom scripts in MATLAB version R2019b (The MathWorks Inc.). Data were filtered using third-order Butterworth filter in 0.001-40 Hz range before estimation of RMS sway.

### Clinical examination and questionnaires

Symptoms were assessed using the Dizziness Handicap Inventory (DHI); and Activities-Specific Balance Confidence scale (ABC). The Addenbrooke’s Cognitive Examination Revised (ACE-R), assessed cognition and the Patient Health Questionnaire 9 (PHQ-9) assessed depression severity. A full clinical examination was performed at 0, 3 and 6months, including assessments of mental state and capacity, eye movements (including the VOR), Romberg, gait, tandem stance, tandem gait, limb tone, power and coordination, and tendon and plantar reflexes, and sensory testing, including pain and joint position.

The patients were initially assessed on the ward by BMS and HMR, both experienced vestibular neurologists who performed a detailed and focused clinical examination including eye movement and vestibular neurological assessment including for signs of peripheral vestibular loss, e.g., doll’s eyes and head impulse manoeuvre, assessing for nystagmus in the primary position with and without visual fixation, and via fundoscopy assessing for any nystagmus with and without visual fixation and assessing for peripheral (e.g. BPPV) and/or central positional nystagmus.

### Statistical analysis of objective and subjective vestibular measures

#### Link between recovery of balance and vestibular agnosia

To assess the mechanistic link between recovery from imbalance and vestibular agnosia, we performed three analyses, hence a corrected *P* < 0.0167 was considered statistically significant. The analyses included: i) a one-way repeated measures ANOVA with factors group (VA+ and VA-) and timepoints (0 and 6 months) using sway RMS as outcome; ii) a correlation was performed between the absolute change in sway and absolute change in VPTs from acute to 6month period; iii) two correlations were performed of absolute change in sway and absolute change in VPTs within VA+ and VA-groups as well. The Bartlett and Levene’s tests were used to assess for the equality of variances, whereas the normalcy of data distribution was assessed using the Shapiro-Wilk test.

As analyses with absolute changes also included patients who were at relatively normal levels, a separate logistic model was used to assess whether vestibular agnosia predicted composite vestibular recovery (balance and vestibular perception). Patients were categorized at 6 months as recovered/normal and non-recovered using the normative balance and vestibular perceptual threshold data from healthy controls with a cut-off of mean + 2SD. This resulted in 12 (of 34) patients who did not make a full recovery to the normative levels. In the model, the patient recovery status was included as a dependent variable whereas the acute vestibular agnosia status (VA+ or VA-) and sex were added as factors. Since our cohort had more males than females (8 of 34 at follow-up), sex was added in the null model to control for its effects.

To supplement the findings from the model, a post hoc analysis is also reported indicating the difference in balance between groups (VA+, VA-, and controls) at 6 months follow-up. Kruskal-Wallis rank sum test (due to unequal variances – Bartlett test) with post hoc Dunn test (Bonferroni adjusted) was used with sway RMS as outcome.

#### Link between subjective symptoms and objective vestibular function

To assess our prediction of lack of concordance between objective and subjective measures, six correlations between objective (balance and VPTs) and subjective measures (DHI, ABC, PHQ-9) were performed, and three correlations within subjective measures (DHI, ABC, PHQ-9) were performed. Thus, a corrected p-value for 9 comparisons (*P* < 0.0055) was used for a correlation to be considered statistically significant. Shapiro-wilk’s test was used to assess the normality assumption and Spearman’s rank correlation was performed if data was not normally distributed.

To assess the link of longitudinal change of symptom scores (DHI) with longitudinal change of objective measures of balance and VPTs in different groups (VA+ and VA-), a one-way repeated measures ANOVA with factors group (VA+ and VA-) and timepoints (0 and 6 months) was performed using DHI score as outcome.

In the manuscript, the term objective vestibular function or recovery is used for the laboratory assessed measures of balance and VPTs, whereas the term subjective symptoms or subjective recovery is used for the subjectively acquired questionnaires (e.g., DHI).

We used R (https://www.R-project.org/, version 4.2.3; 2023-03-15) and JASP^25^ (version 0.17.2) for statistical analyses and graphical outputs.

### Neuroimaging

#### Image acquisition

Structural and functional MRI images were acquired at time 0 and 6 months using a 3 T Siemens Verio MRI scanner using a 32-channel head coil. The details of scanning protocol is reported in supplementary whereas preprocessing and analysis pipelines for diffusion tensor imaging analysis, and VBM analysis are reported below.

### Structural brain imaging – diffusion tensor imaging (DTI)

#### Pre-processing

Diffusion-weighted images were processed following the standard tract based spatial statistics (TBSS)^26^ pipeline in the FMRIB Software Library (version 5.0.8),^27^ including correcting susceptibility and eddy current induced distortions and diffusion tensor fitting. Brain extraction was performed using HD-BET.^28^ Tensor-based registration was performed using DTI-TK,^29^ and involved the creation of a group template using affine and non-linear diffeomorphic registrations followed by registration of participant diffusion imaging to the template. Images were warped to 1mm isotropic space, and the mean FA map produced was thresholded at 0.2 to produce a white matter skeleton. Subject FA data was then projected onto the mean FA skeleton. Each participant’s longitudinal change in FA was calculated with ‘fslmaths’ using the skeletonised images and the images were then merged into a single 4D image and used for group-level voxelwise analysis.

#### Group level analysis

Normalized change values (z-scores) of both behavioural measures, vestibular perceptual thresholds (VPT) and the RMS sway in soft surface-eyes closed condition, were used as covariates to identify the interaction of recovery of vestibular perception and postural balance as well as their respective main effects in FSL.^27^

#### Statistical Analysis

All group level FA changes were evaluated using non-parametric permutation statistics. Threshold-free cluster enhancement (TFCE)^30^ at *P* < 0.05 was used for multiple comparison correction. Findings are reported after correction for 6 contrasts at *P* < 0.0083 i.e., positive and negative correlation (equivalent to two one-sided t-tests) for each main effect (change in sway & change in vestibular perceptual thresholds), and for the interaction of the same measures.

### Structural brain imaging – voxel-based morphometry (VBM)

#### Pre-processing

Data were pre-processed using the CAT12 Toolbox^31^ using voxel-based morphometry (VBM) analysis^32^ with SPM12 (http://www.fil.ion.ucl.ac.uk/spm/). The CAT12 automated preprocessing steps included spatial-adaptive non-local means denoising,^33^ and interpolation. Data were then bias corrected, affine-registered, and segmented using unified segmentation approach allowing skull-stripping, and the intensity normalized (to control for hyperintensities), and re-segmented using adaptive maximum a posteriori.^34^ The segmentation was controlled for partial volume.^35^ The data were transformed to MNI space and smoothed using a gaussian kernel with FWHM of 6mm.^31^

#### Group level analysis

For the group level analysis, normalized change values of vestibular perceptual thresholds (VPT) and the RMS sway, were used as covariates to identify the interaction of recovery of vestibular perception and postural balance as well as their respective main effects in SPM. Total intracranial volume (TIV) was not included as a covariate since the analysis focuses on within subject changes for which TIV remains constant for all timepoints.^31^

#### Statistical Analysis

The GM images were analysed in group comparison using a flexible factorial design, which accounts for the longitudinal nature for each subject’s data.^31^ Findings are reported after correcting for multiple comparisons at cluster-level using family-wise error (FWE) correction at *P* < 0.05. Findings were also reported after correcting for the statistical evaluation of 3 contrasts i.e., two main effects (vestibular perceptual thresholds and RMS Sway) and their interaction (corrected *P* = 0.016). Cluster height threshold for grey-matter specific VBM analysis was estimated to be F = 39.46 with an extent threshold of k = 10 voxels. The findings were shown using “BrainNet Viewer”,^36^ and in MNI coordinates.

## Results

### Recruitment and demographics

#### Participants

Recruitment and follow-up testing took place between September 2017 and September 2020. N = 39 patients were recruited acutely. N = 34 completed behavioural testing at 6 months, except one patient at 3 months and six at 12 months (primarily due to the COVID19 pandemic). Numbers obtained for longitudinal imaging data were as follows: N = 33 patients for VBM, N = 27 for the DTI (6 dropouts). Resting-state fMRI analysis from N = 17 (of N = 27) patients is separately reported in supplementary file due to a limited sample (N = 11 patients were removed either due to mismatched field of view parameters or outlier scans). Demographic details of this subsample (N = 17) are also provided in supplementary Table S2.

### 37 age and sex matched healthy controls (Age: 40.78 ± 14.75 | Mean ± SD; 21 Females) completed behavioural and neuroimaging testing once

#### Demographics

The average age of the cohort (Table 1) was 41.64 years (standard deviation: 13 y, range: 18 – 65 years) with 71.80% male (28 M; 11 F). The commonest cause of injury was falls recorded in 21 patients (of 39). The second commonest cause was road traffic accident (RTA) recorded in 15 patients (of 39). According to the Mayo TBI severity criteria,^37^ 35 of 39 patients were moderate-to-severe with 4 mild-probable TBIs.

**Table 1.** Participants demographics and clinical details of injury, clinical examination and reported history at each assessment.

| Subject | MOI | Severity<br>MAYO | PTA | BPPV |  |  | Headache<br>0 month |
| --- | --- | --- | --- | --- | --- | --- | --- |
|  |  |  |  | 0 month | 3 month | 6 month |  |
| 1 | RTA | Mod-Sev | 1 | + | - | - | - |
| 2 | RTA | Mod-Sev | 1 | + | + | + | + |
| 3 | Fall | Mod-Sev | 1 | + | - | NA | - |
| 4 | RTA | Mod-Sev | 0 | - | - | - | - |
| 5 | RTA | Mild-Prob | 0 | + | NA | NA | - |
| 6 | Fall | Mod-Sev | 1 | - | - | - | - |
| 7 | RTA | Mild-Prob | 0 | - | - | - | - |
| 8 | Assault | Mod-Sev | 0 | - | NA | NA | + |
| 9 | RTA | Mod-Sev | 0 | - | - | - | + |
| 10 | Fall | Mod-Sev | 0 | + | - | + | - |
| 11 | Fall | Mod-Sev | 1 | - | - | - | - |
| 12 | RTA | Mod-Sev | 1 | - | - | - | - |
| 13 | Fall | Mild-Prob | 0 | + | - | - | + |
| 14 | Fall | Mod-Sev | 0 | + | - | - | - |
| 15 | RTA | Mod-Sev | 0 | + | NA | + | - |
| 16 | Fall | Mod-Sev | 0 | - | NA | NA | - |
| 17 | Fall | Mod-Sev | 0 | + | - | - | - |
| 18 | RTA | Mod-Sev | 0 | - | - | - | + |
| 19 | Fall | Mod-Sev | 0 | + | NA | - | - |
| 20 | Fall | Mod-Sev | 0 | - | - | - | + |
| 21 | Fall | Mod-Sev | 0 | - | - | - | + |
| 22 | Fall | Mod-Sev | 0 | - | + | - | - |
| 23 | Fall | Mod-Sev | 1 | + | - | + | + |
| 24 | RTA | Mod-Sev | 1 | - | NA | + | - |
| 25 | Fall | Mod-Sev | 1 | + | NA | - | - |
| 26 | Fall | Mod-Sev | 1 | + | NA | NA | - |
| 27 | Assault | Mod-Sev | 1 | - | + | - | - |
| 28 | RTA | Mod-Sev | 1 | - | NA | NA | + |
| 29 | Fall | Mod-Sev | 0 | - | NA | - | - |
| 30 | Fall | Mod-Sev | 0 | + | - | - | - |
| 31 | Fall | Mod-Sev | 0 | + | + | + | - |
| 32 | RTA | Mod-Sev | 1 | - | - | - | - |
| 33 | Assault | Mod-Sev | 1 | - | NA | - | + |
| 34 | Fall | Mod-Sev | 1 | + | NA | - | + |
| 35 | Fall | Mild-Prob | 0 | + | - | - | + |
| 36 | RTA | Mod-Sev | 1 | + | NA | - | - |
| 37 | RTA | Mod-Sev | 1 | - | - | - | - |
| 38 | Fall | Mod-Sev | 1 | + | NA | - | - |
| 39 | RTA | Mod-Sev | 1 | - | NA | - | - |

NA: not available; +/1: present; -/0: absent; MOI: mode of injury; Mod-Sev: moderate to severe; RTA: road traffic accident

#### Clinical recovery of vestibular function

##### Resolution and recurrence of BPPV (benign paroxysmal positional vertigo)

Acutely, 19 of 39 (48.7%) patients had BPPV, and all of whom received repositioning manoeuvres and were clear of BPPV at the time of testing at time 0. We saw 24 patients (of 39) for 3-month assessment, 4 of 24 patients had BPPV at 3 months, of whom 2 were ‘new onset’ BPPV at 3 months (i.e. BPPV not observed at 0 months despite ward review by senior neurologists BMS and/or HMR); indicating a change in BPPV prevalence from 48.7% at time 0 months (19 of 39 patients) to 16.67% (4 of 24 patients) at 3 months. At 6 months, we assessed 33 patients of whom 6 patients had BPPV representing a 31.6% recurrence rate despite apparent previous treatment to resolution by repositioning manoeuvres. Note that at each testing session, patients with recurrent (or new) BPPV at follow-up, were treated until resolution, returning a few days later for testing.

In the 146 patients we examined acutely for this study, we found that cases with skull fracture were more likely to have BPPV than those without fractures, indicating a force dependency of BPPV.^23^

##### Recovery of subjective dizziness and perceived imbalance does not link to objective recovery

The DHI (subjective dizziness and balance) scores generally reduced at follow-up (Fig 2A & Fig 3E) except a few patients with VA+ whose DHI scores increased (Fig 3E). Notably, change in DHI did not correlate with change in vestibular perceptual thresholds (VPTs) or change in sway (Table 2). Similarly, change in subjective balance (ABC scale), did not corelate with change in objectively assessed balance or with VPTs (Table 2). Thus, subjective vestibular symptom scores did not correlate with their corresponding objective measures.

**Figure 2.**
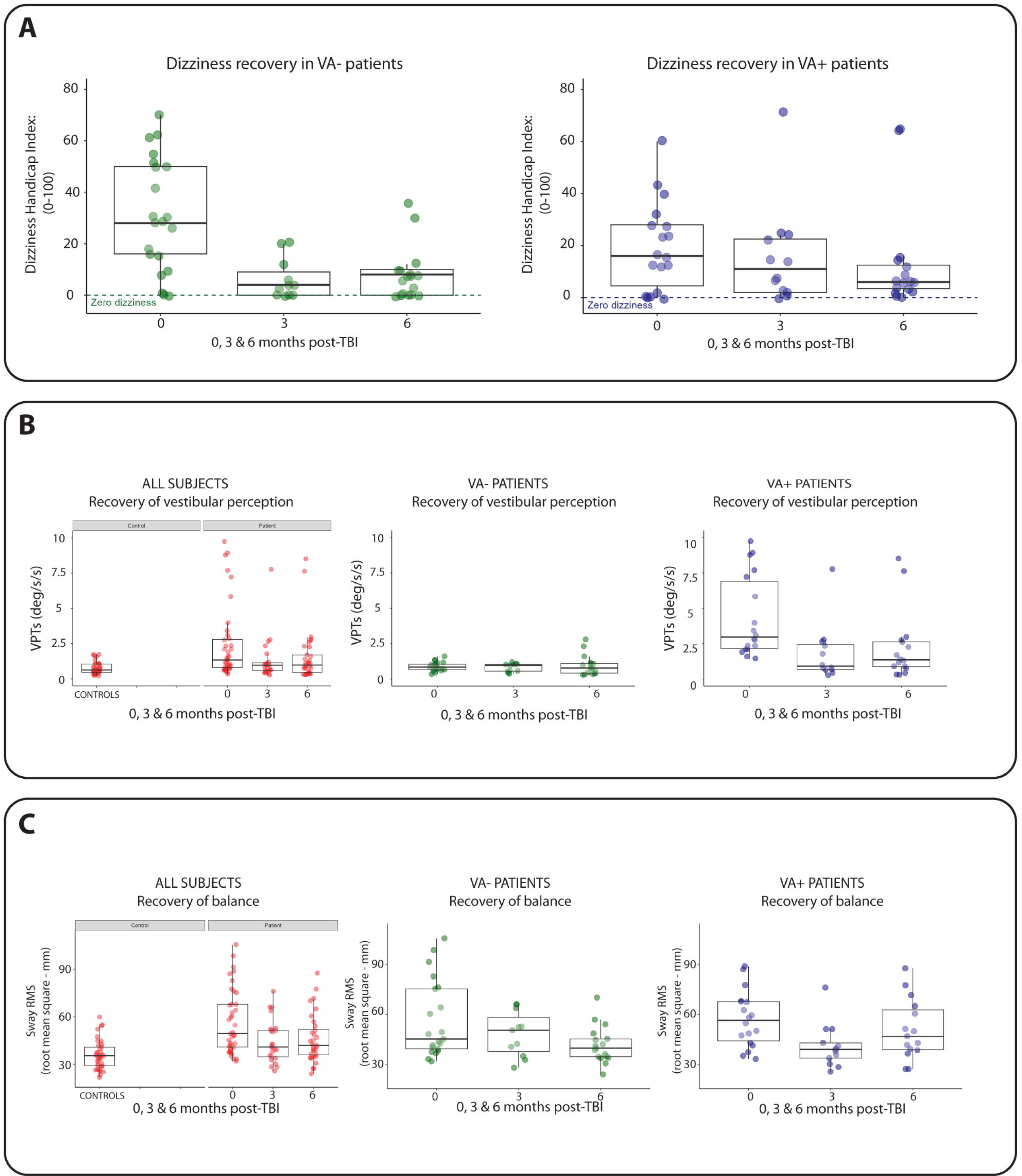
Recovery of dizziness symptoms, vestibular perceptual thresholds of self-motion and balance. (A) Recovery of dizziness over time. The dizziness handicap inventory (DHI) overall improved for both patients without vestibular agnosia (VA-; Fig 2A, shown in green) and patients with vestibular agnosia (VA+; Fig 2A, shown in blue). **(B) Recovery of objective vestibular perceptual function over time.** Overall vestibular perceptual thresholds (VPTs) improved – i.e. reduced – over time across all patients (Fig 2B – left panel, in red). For VA-patients, VPTs remained mainly within the normal range (Fig 2B – middle panel, in green). For VA+ patients, there was persistence of abnormal VPTs at follow-up (Fig 2B – right panel, in blue). **(C) Recovery of objective balance function.** Sway is shown as root mean square (RMS) of sway obtained during ‘eyes closed with soft surface’ condition. Overall sway improved – i.e. reduced – over time for all patients (Fig 2C – left panel, in red). Balance recovery was worse however in the VA+ group with several patients showing persistently elevated sway RMS above the control range (Fig 2C – right panel, in blue).

**Figure 3.**
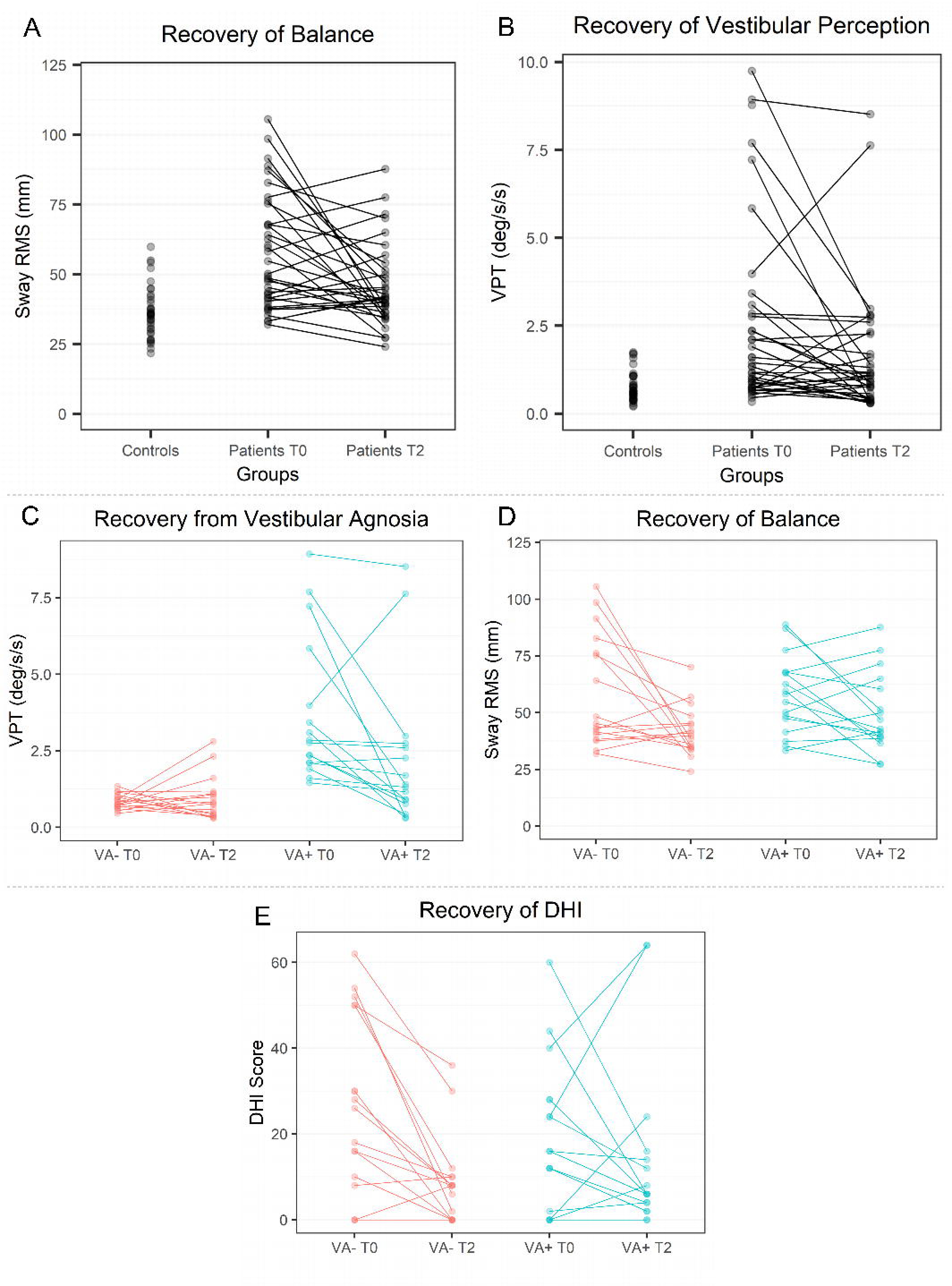
Longitudinal trajectories of recovery of balance, vestibular perceptual thresholds of self-motion, and dizziness symptoms. (A-B) Recovery of patients’ balance and vestibular perceptual thresholds compared to controls. Patients’ vestibular perceptual thresholds (VPTs) and balance were higher than controls acutely (T0) and generally reduced on 6-month follow-up (T2). **(C) Recovery of vestibular perceptual thresholds.** Overall VPTs improved from acute (T0) to follow-up (T2) in patients with VA+. Few patients with VA+ got worse on 6-month follow-up (T2) (in blue) whereas few VA-patients also developed VA (in red). **(D) Recovery of objective balance function.** Sway is shown as root mean square (RMS) of sway obtained during ‘eyes closed with soft surface’ condition. Overall sway improved – i.e., reduced – over time for VA-patients (in red). However, several VA+ had persistent imbalance at 6-month follow-up (T2) and some also got worse (in blue). **(E) Recovery of dizziness over time.** Dizziness scores (via “Dizziness Handicap Inventory”) generally reduced for all patients except a few patients with VA+ who got worse from acute (T0) to 6-month (T2) follow-up (in blue).

**Table 2.** Correlations of subjective questionnaires and objective measures.

| Measure 1 | Measure 2 | Spearman's rho | P - value | 95% Confidence Interval |
| --- | --- | --- | --- | --- |
| DHI | Balance<sup>a</sup> | 0.135 | 0.447 | [-0.209, 0.475] |
| ABC | Balance<sup>a</sup> | -0.234 | 0.191 | [-0.573, 0.134] |
| PHQ-9 | Balance<sup>a</sup> | 0.347 | *0.048 | [-0.032, 0.628] |
| DHI | VPT | 0.092 | 0.603 | [-0.307, 0.450] |
| ABC | VPT | -0.298 | 0.093 | [-0.605, 0.055] |
| PHQ-9 | VPT | 0.281 | 0.113 | [-0.127, 0.595] |
| DHI | ABC | -0.507 | **0.003 | [-0.764, -0.157] |
| DHI | PHQ-9 | 0.732 | **$1.27 \times 10^{-6}$ | [0.450, 0.899] |
| ABC | PHQ-9 | -0.550 | **0.001 | [-0.768, -0.245] |

DHI: Dizziness Handicap inventory; ABC: Activities-Specific Balance Confidence scale; PHQ-9: Patient Health Questionnaire-9; VPT: vestibular perceptual thresholds.
\*: statistically significant at $P < 0.05$
\*\*: statistically significant at corrected p-value for 9 correlations ( $P < 0.0055$ )
<sup>a</sup>: Balance measured as root-mean square sway

In contrast, the change in subjective questionnaires (DHI, ABC, & PHQ-9) over follow-up were significantly correlated with each other (corrected for multiple comparisons; Table 2). It follows that symptomatic ‘dizziness’ cannot be used on its own as a proxy for vestibular recovery post-TBI.

Fig 2A and Fig 3E shows the difference in dizziness recovery when divided into patients with and without vestibular agnosia. Patients with VA (VA+) have lower acute dizziness mean score (Fig 2A) despite having significantly worse acute clinical deficit.^7^ An interaction of vestibular agnosia status (VA+ or VA-) and timepoints (acute and 6 month) with DHI scores as outcome, indicated a borderline statistical difference in symptomatic recovery trajectories of VA subgroups (F(1,32) = 4.174, *P* = 0.049).

##### Patients’ balance recovery and its link to vestibular agnosia

In general, sway improved (reduced) over time for all patients (Fig 2C & Fig 3A), however, balance recovery was worse in the VA+ group with several patients showing persistently elevated sway RMS or further worsening (increase) of sway (Fig 3D). Similarly, vestibular perceptual thresholds (VPTs) generally improved (reduced) from acute to follow-up (Fig 2B & Fig 3B). When we looked at the recovery of vestibular perceptual thresholds (VPTs) split into patients with acute VA+ vs. VA-(Fig 2B and Fig 3C), we noted that acute VA+ patients showed higher rates of persisting VA+ at 6-12 month follow-up. We also noted that two acute VA-patients developed vestibular agnosia at the 6-month follow-up (Fig 3C), one of whom also developed imbalance despite having normal balance acutely.

Using the ‘eyes-closed-soft-surface’ balance condition (which we previously found to be the best discriminator of imbalance of patients (VA+ and VA-) from controls in acute TBI),^7^ a within patient one-way repeated measures ANOVA with factors “Group” (VA+ vs. VA-) and “Time” (0 vs. 6 months) did not show a significant interaction F(1,32) = 1.251 (*P* = 0.272, η_p_^2^ = 0.038).

Correlating the change of VPTs and change of sway from acute to follow-up (Fig 4A), showed a positive correlation between change of VPTs and sway (ρ^2^ = 0.23, *P* = 0.0043; corrected *P* < 0.0167). Since correlation of all patients could mask the changes occurring at sub-group level, we then looked at the link between change of VPTs and change of sway from acute to follow-up by stratifying patients into VA+ and VA- and performing separate correlations. Fig 4B shows that the correlation between balance and VPT change is more robust for the VA+ group (corrected *P* < 0.0167) than for the VA-group. However, the statistical difference between the two correlations is not significant (*P* > 0.05, 95% CI [-0.60 0.35]).

**Figure 4.**
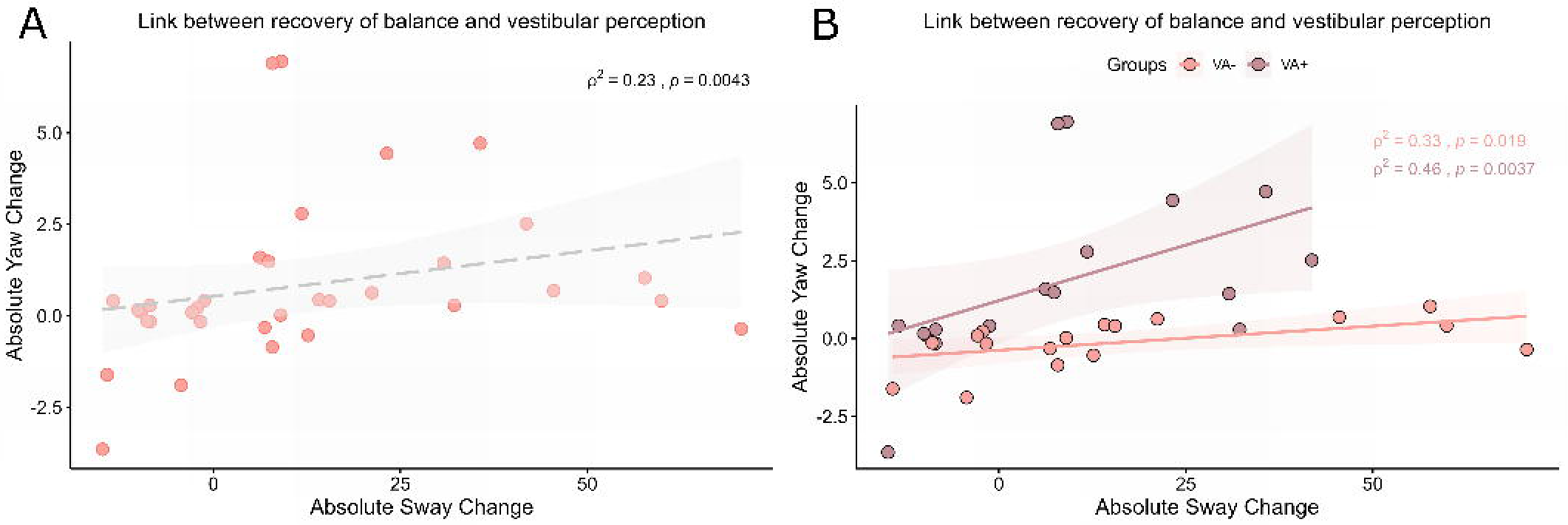
Relationship between recovery from vestibular agnosia and balance. (A) Correlation between continuous measures of vestibular perception and balance. Change in sway RMS and VPTs from acute to 6-month follow up testing were linked. **(B) Recovery of balance linked to VA recovery according to acute VA status.** Longitudinal change in the balance-VA relationship appeared more robust in VA+ vs VA-patients (although the two correlations were not statistically different). (RMS: root mean square; VPTs: vestibular perceptual thresholds; VA+: group with vestibular agnosia; VA-: group without vestibular agnosia)

As the above analyses using absolute VPTs and sway changes included all patients, including those with relatively normal balance, we used a separate logistic model to assess if VA is a predictor of vestibular recovery (i.e., of balance and perception) in TBI (*see methods for classification details of recovery vs non-recovery patients*). The model indicated that VA indeed is a marker of worse recovery of balance and vestibular perception. (Model summary: χ^2^(31) = 5.895, *P* = 0.015). Model coefficients are reported in Table 3. To supplement the model’s conclusion, a post-hoc analysis to assess the difference in sway at 6 months between VA+ and VA-in comparison to healthy controls indicated that VA+ (*P* = 0.0043), but not VA-patients (*P* > 0.05), had worse balance compared to controls (χ^2^ = 10.69; *P* = 0.0048; where VA+ vs. VA-was not statistically different when corrected (*P* > 0.05)).

**Table 3.** Logistic model predicting vestibular recovery.

|  | Estimate | Standard Error | Odds Ratio | z | Wald Statistic | df | p | Confidence Interval |
| --- | --- | --- | --- | --- | --- | --- | --- | --- |
| (Intercept) | 0.702 | 0.835 | 2.017 | 0.840 | 3.937 | 1 | 0.047 | [0.39 10.37] |
| VA (1) | -0.659 | 1.053 | 0.517 | -0.626 | 0.392 | 1 | 0.531 | [0.02 0.81] |
| Gender (1) | 4.585 | 2.092 | 98.007 | 2.192 | 4.806 | 1 | 0.028 | [0.57 27.34] |

VA: vestibular agnosia. Confidence interval for odds ratio.

#### Neuroimaging correlates of vestibular recovery

##### Brain white matter microstructural (‘DTI’) connectivity and link to vestibular recovery

A significant interaction of longitudinal change (Δ) of sway and of VPTs (vestibular perceptual thresholds) with longitudinal change (Δ) in FA was localised primarily in posterior corpus callosal regions including the splenium of the corpus callosum, forceps major, and body of the corpus callosum (TFCE corrected findings at *P* < 0.0083; Fig 5A & Table 4). No main effects (of Δ sway or Δ VPTs) were found.

**Figure 5.**
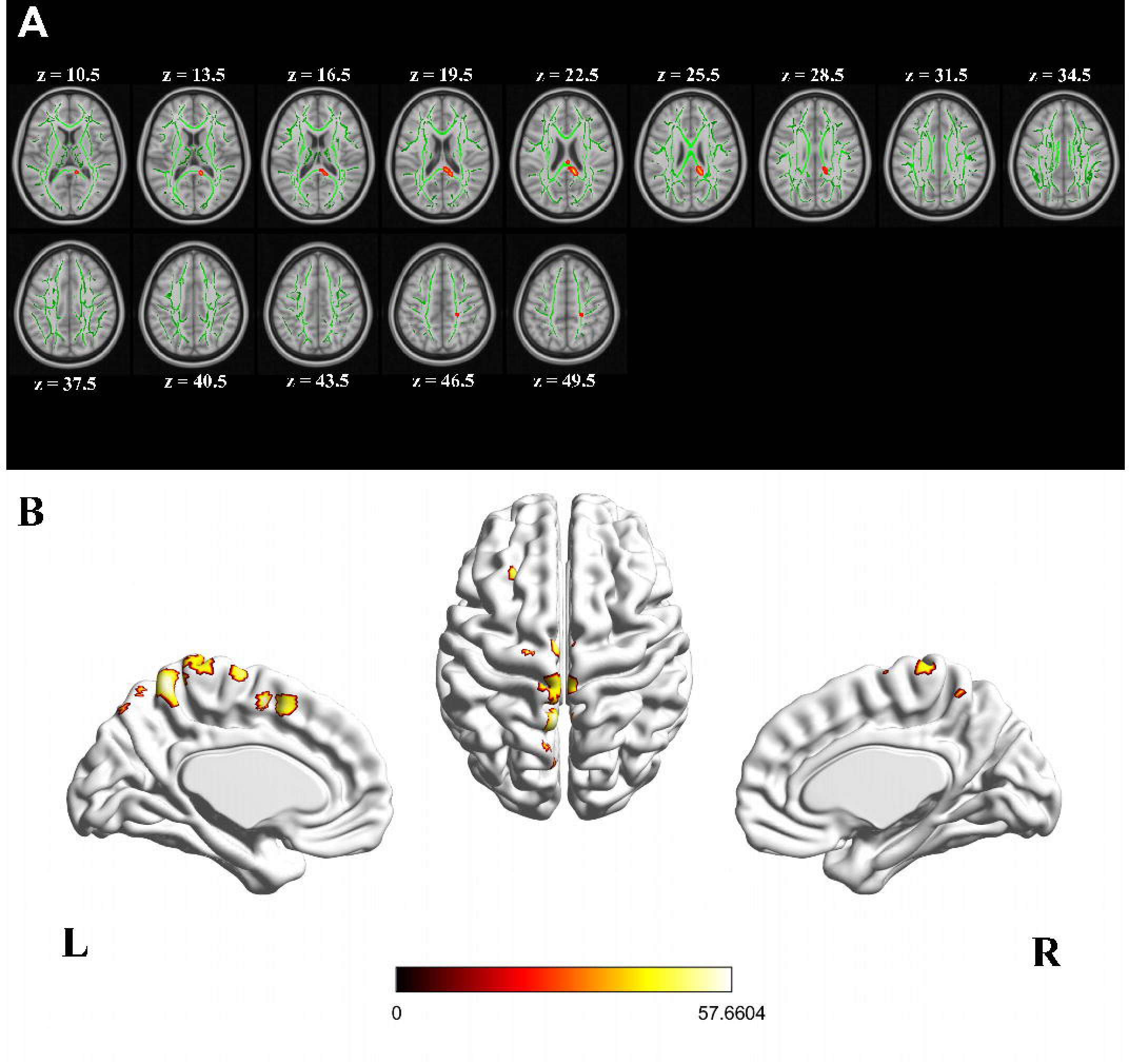
Interaction of change (Δ) in vestibular perceptual thresholds (VPTs), Δ sway, and the Δ connectivity values from different imaging modalities. L and R represent left and right hemisphere convention for all panels (A-B). **(A)** Diffusion tensor imaging analysis indicating a significant interaction (Δ VPT × Δ sway × Δ FA). Significant regions are highlighted in red and overlayed on mean FA skeleton (green) of all participants. (TFCE corrected findings at *P* < 0.0083). **(B)** Results from voxel-based morphometry (VBM) analysis showing interaction (Δ VPT × Δ sway × Δ Volume) at two clusters centred at left supplementary motor cortex, two at left precuneus, one at left precentral gyrus, one at left midfrontal gyrus, and one at precentral lobule (FWE corrected at P < 0.05). (*Colour-bar indicate F statistic)*.

**Table 4.**
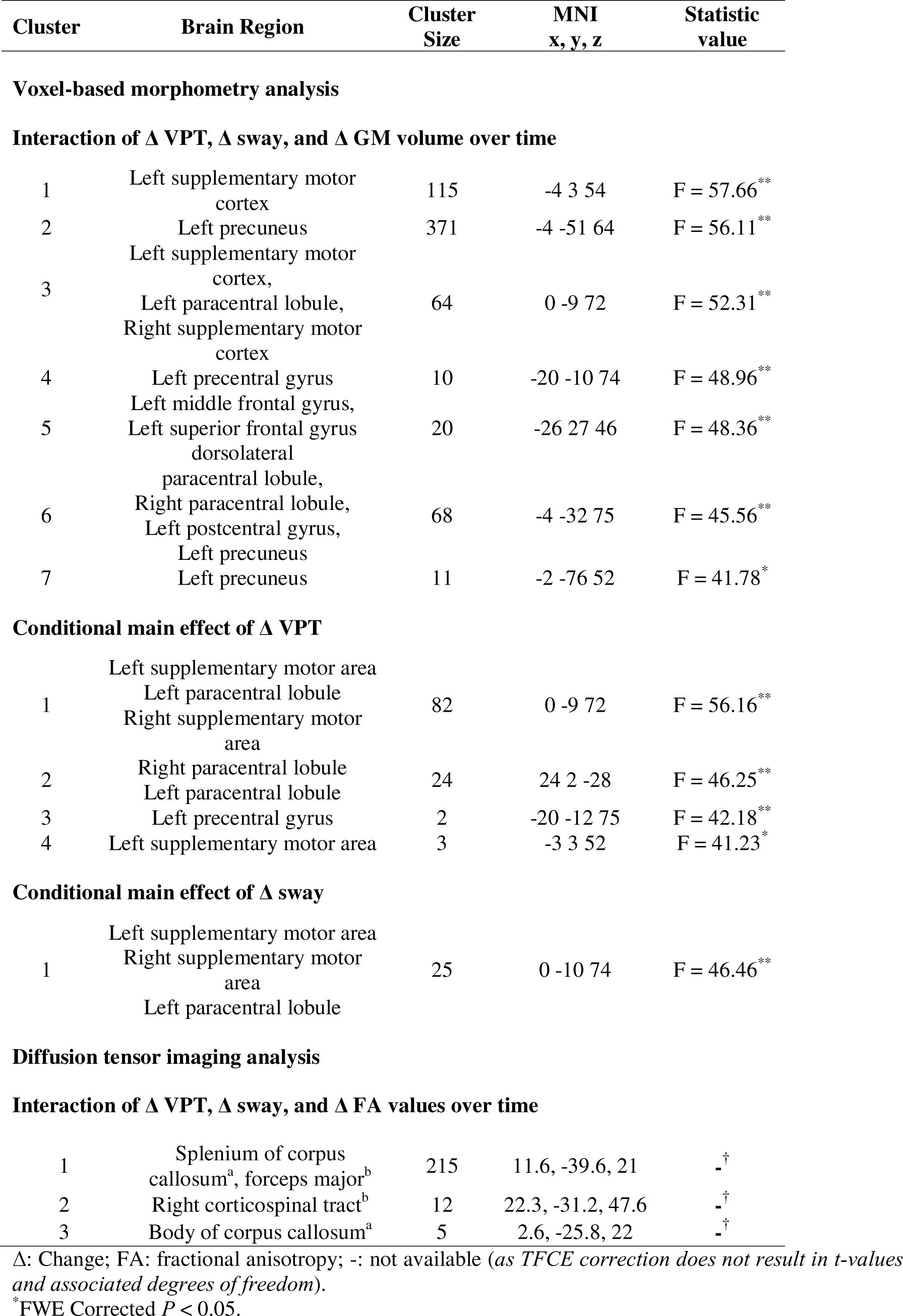
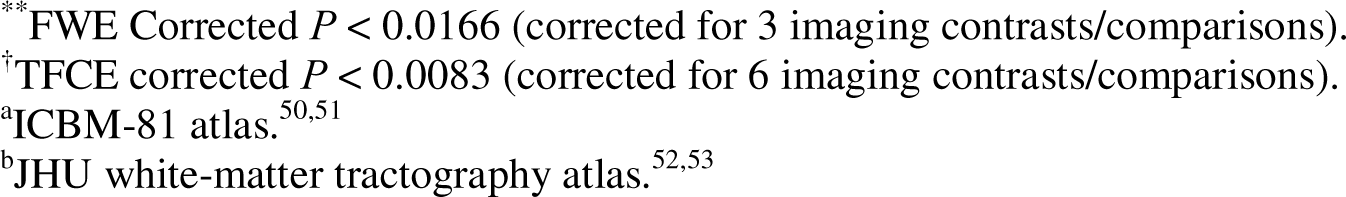
Interaction and main effects of Δ VPT, Δ sway, and Δ GM volume over time.

##### Brain grey-matter volume correlates of behavioural change (balance and vestibular perceptual thresholds)

A significant interaction of the behavioural measures (Δ sway and Δ vestibular perceptual thresholds (VPTs)) with longitudinal change in grey-matter volume (Δ GM volume) was found primarily in the left hemisphere (Fig 5B; FWE corrected *P* < 0.05), including two clusters in the left SMA, two in the left precuneus, and single clusters in the left mid-frontal gyrus, left precentral gyrus, and the paracentral lobule (Table 4; corrected for three comparisons *P* < 0.0166).

## Discussion

In this first acute-prospective-longitudinal study of vestibular function in acute hospitalised moderate-severe TBI patients, we show for the first time that acute vestibular agnosia (VA) on its own, predicts worse vestibular recovery (of balance and perception) at 6 months. Critically, change in subjective vestibular symptom scales (DHI and ABC) did not predict objective vestibular recovery. Regarding underlying brain mechanisms, our structural (white and grey matter) brain imaging found that vestibular recovery was mediated primarily by recovery of posterior corpus callosal structural integrity, mediating sensory signalling, and motor and premotor cortical regions, mediating efferent signalling. These data thus support the notion that vestibular cortical circuits mediating the vestibular signals of head motion are highly distributed,^7,22,38–41^ with recovery of interhemispheric sensori-motor signalling being critical for post-TBI recovery of vestibular-mediated balance and perception.

### Vestibular recovery and subjective ‘dizziness’ symptom scores

Within-subject acute and follow-up balance measures indicated that generally patients’ sway improved (decreased) over time however several patients with vestibular agnosia either did not improve sway performance or got worse (Fig 3D). We also found that the recovery from imbalance and vestibular agnosia was linked such that acute VA status resulted in worse balance recovery at 6 months. While the occurrence of vestibular agnosia has previously been reported in several studies in elderly with imbalance,^10–15^ none established the clinical relevance of vestibular agnosia for the prognosis of such individuals. Our current report in TBI is the first to confirm worse prognostic outcome for unbalanced individuals with vestibular agnosia.

Notably, in the previous reports in elderly,^12,13,15^ the individuals reported having postural unsteadiness or postural instability; and while authors argued that vestibular agnosia could result in lack of awareness about their unsteadiness, no findings were reported. Contrary to previous reports, we show a lack of concordance between objectively assessed balance and subjectively assessed balance using symptom scales indicating patients’ lack of awareness about their postural instability. This implies that subjective scores are a proxy for patients’ symptomatic well-being, but they do not inform upon objective recovery and hence poorly track brain injury-related recovery. It follows that symptomatic ‘dizziness’ cannot be used on its own as a proxy for vestibular recovery post-TBI.

Finally, few patients who did not have acute VA, developed VA (i.e., elevated vestibular perceptual thresholds) at 6-12 month follow-up. We speculate that persistence or progression of VA could be linked to falls, either via undiagnosed vestibular conditions (e.g., BPPV) or by the mechanistic overlap between VA and imbalance. Current studies tracking cognitive function and falls over time in TBI survivors, should consider tracking VA since progressive changes in VA could herald the onset of neurodegeneration and falls, both of which are linked to long term survival in TBI survivors.^21^

### The brain mechanisms mediating recovery of vestibular function

The data linking vestibular perception and balance are sparse, however the link was first theorised to occur based upon the clinical observation of vestibular agnosia in an elderly patient with diffuse white matter small vessel disease, falls, and BPPV without vertigo.^42^ In a subsequent study we failed to find any effect of acute focal stroke upon vestibular perceptual thresholds,^39^ implying that VA, and potentially it’s link to balance, was mediated by brain networks^38^ rather than a discrete brain region. In this study, we refine our understanding of the brain networks supporting VA and linked imbalance and for the first time, the neuroimaging correlates of their recovery.

Longitudinal changes in white matter microstructure (via DTI) of the splenium of the posterior corpus callosum were linked with the recovery of imbalance and vestibular agnosia. Indeed, interhemispheric disconnection in the genu, body, & splenium of corpus callosum was confirmed in unbalanced TBI patients compared to healthy controls.^7^ More convincing however, is that TBI patients with imbalance had worse damage to the genu of the corpus callosum than TBI patients with normal balance.^7^ These findings are congruent with that of our longitudinal fMRI data (supplementary) and with our previous studies,^7,22^ and buttresses the notion that interhemispheric disconnection in TBI leads to vestibular dysfunction with imbalance and vestibular agnosia.

Our longitudinal VBM findings link the recovery from imbalance and vestibular agnosia with volumetric change in left precuneus in the posterior parietal cortex (PPC), which has projections to an important vestibular processing region, the parieto-insular vestibular cortex (PIVC)^43^, which previous studies have shown is transcallosally connected via the splenium.^44^ That our VBM findings were primarily in posterior cortical regions, again supports the notion that impaired vestibular perception and balance in acute TBI is an acute disconnection syndrome, and their recovery is correspondingly mediated by recovery of posterior interhemispheric connectivity.

Our structural analysis (DTI and VBM) also showed that vestibular recovery is also linked to recovery of primary and secondary motor cortical areas and their connections, including the corticospinal tract (Table 4). Frontal cortical regions have been linked to short-term balance training^45^ and long-term training in elite dancers.^38^ Apart from damage to the corticospinal tract motor output to the spinal motor neurones, injury to the extensive recurrent projections between primary and secondary motor areas^46^, explains how motor cortical damage impairs efferent control of balance in TBI.^47^

### Limitations

The patient cohort was purposely highly refined by strict inclusion and exclusion criteria and limited to a relatively young age (cohort average age 41.6 years) to minimise premorbid brain (e.g. dementia), psychiatric or medical diseases (e.g. diabetes mellitus), which could confound the recovery of these patients. The 15% of hospitalized TBI cases with acute unilateral peripheral hypofunction^3^ were excluded since this can complicate assessing vestibular ocular and perceptual vestibular thresholds^48^. Our findings are thus primarily related to brain injury, but poorly generalize to the elderly, patients with peripheral vestibular injury, and those with medical and psychiatric comorbidities. Assessing factors predicting recovery in multi-morbid patients is a necessary next step, and we predict vestibular recovery in older, multi-morbid patients with TBI will be significantly worse.

Our study is of relatively small size and is linked to our strict exclusion criteria, despite screening c. 1000 patients, and the detailed acute and prospective assessment of each patient. Our sample size is however comparable to similar mechanistic studies,^38,49^ although these studies were cross-sectional.

Finally, patients were recruited on the basis of a TBI in unbiased fashion and not because they were dizzy and/or unbalanced. The frequencies of vestibular dysfunction thus reflect the heavy burden of vestibular dysfunction in acute TBI. Excluding cases with peripheral vestibular loss likely excluded cases with even worse imbalance when central imbalance is compounded by peripheral loss.

### Conclusion

In this first acute-prospective study assessing recovery of objective vestibular function, we found that vestibular agnosia results in worse balance recovery in unbalanced patients. Moreover, vestibular agnosia and imbalance recovery is mediated by recovery of interhemispheric structural and functional connectivity. Patients with VA are at increased falls risk from: (i) damage to overlapping brain circuits mediating balance and vestibular perception of self-motion; (ii) and lack of vertigo symptoms resulting in missed treatable vestibular conditions. Given that subjective symptoms of dizziness unreliably indicate brain injury severity, its recovery, and treatable vestibular diagnoses, one implication (albeit requiring additional study evidence) is that all TBI patients require detailed vestibular neurological assessment regardless of symptoms. Our finding of progressive worsening of balance and vestibular perception in some individuals requires replication in larger, long-term follow-up studies, important if this represents progressive neurodegeneration. TBI survivors, like other vulnerable groups (elderly,^18^ dementia) should be screened for common treatable balance conditions (e.g. BPPV) irrespective of vertigo symptoms, in order to at least partially modify falls risk and long-term mortality from falls. Finally, interventions enhancing bihemispheric connectivity (druggable and non-druggable), should be considered for improving post-TBI vestibular recovery.

## Acknowledgements

We are grateful to our patients and healthy control volunteers for their participation. The support of their families and friends was critical in the recruitment and testing process. We are grateful for the Headway Charity for discussions about the project and its impact upon patients and their support network. We are also grateful to the major trauma ward teams at St Mary’s Hospital and King’s College Hospital London for their help with recruitment and assessment. We are also grateful for the infrastructure support for this research from the NIHR Imperial Biomedical Research Centre (BRC). We are also very grateful to The Imperial Health Charity who provided important kickstarter funding, including for a clinical fellowship that supported Dr Heiko Rust, who tirelessly and expertly reviewed in-patients and without whom we would not have been able to deliver this project. We look forward to delivering future studies to improve the care of TBI patients.

## Authorship contribution statement

**ZH:** investigation, methodology, analysis, visualization, writing – original draft with BMS. **MM:** investigation, methodology, analysis, visualization, writing – review & editing. **EC:** investigation, methodology, writing – review & editing. **MC:** investigation, methodology, writing – review & editing. **KAZ:** analysis, writing – review & editing. **VT:** investigation, methodology, writing – review & editing. **RMS:** investigation, methodology, writing – review & editing. **HMR:** investigation, methodology, writing – review & editing. **DJS:** Supervision, writing – review & editing. **BMS:** conceptualization, investigation, project administration, funding acquisition, resources, supervision, writing – original draft with ZH.

## Data availability statement

Raw data that support the findings of this study are available from the principal investigator, upon reasonable request. The request would require a formal data sharing agreement, approval from the requesting researcher’s local ethics committee, a formal project outline, and discussion regarding authorship on any research output from the shared data if applicable.

## References

1. Langlois JA, Rutland-Brown W, Wald MM. The epidemiology and impact of traumatic brain injury: A brief overview. J Head Trauma Rehabil. 2006;21(5):375–378. doi:10.1097/00001199-200609000-00001

2. Thomas KE, Stevens JA, Sarmiento K, Wald MM. Fall-related traumatic brain injury deaths and hospitalizations among older adults--United States, 2005. J Safety Res. 2008;39(3):269-272. doi:10.1016/J.JSR.2008.05.001

3. Marcus HJ, Paine H, Sargeant M, et al. Vestibular dysfunction in acute traumatic brain injury. J Neurol. Published online October 1, 2019. doi:10.1007/s00415-019-09403-z

4. Maskell F, Chiarelli P, Isles R. Dizziness after traumatic brain injury: results from an interview study. Brain Inj. 2007;21(7):741–752. doi:10.1080/02699050701472109

5. Maskell F, Chiarelli P, Isles R. Dizziness after traumatic brain injury: Overview and measurement in the clinical setting. Brain Inj. 2006;20(3):293–305. doi:10.1080/02699050500488041

6. Hadi Z, Mahmud M, Seemungal BM. Brain mechanisms explaining postural imbalance in traumatic brain injury: a systematic review. medRxiv. Published online July 16, 2023. doi:10.1101/2023.07.15.23292709

7. Calzolari E, Chepisheva M, Smith RM, et al. Vestibular agnosia in traumatic brain injury and its link to imbalance. Brain. 2021;144(1):128–143. doi:10.1093/brain/awaa386

8. Sargeant M, Sykes E, Saviour M, et al. The utility of the Sports Concussion Assessment Tool in hospitalized traumatic brain injury patients. J Concussion. 2018;2. doi:10.1177/2059700218808121

9. Glaser MA. XXXV The cause of dizziness in head injuries: A vestibular test study in sixty-six patients. Ann Otol Rhinol Laryngol. 1937;46(2):387–408. doi:10.1177/000348943704600209/ASSET/000348943704600209.FP.PNG_V03

10. Seemungal BM, Gunaratne IA, Fleming IO, Gresty MA, Bronstein AM. Perceptual and nystagmic thresholds of vestibular function in yaw. J Vestib Res. 2004;14(6):461–466.

11. Imbaud Genieys S. Vertigo, dizziness and falls in the elderly. Ann d’Oto-Laryngologie Chir Cervico-Faciale. 2007;124(4):189–196. doi:10.1016/j.aorl.2007.04.003

12. Chiarovano E, Vidal PP, Magnani C, Lamas G, Curthoys IS, de Waele C. Absence of rotation perception during warm water caloric irrigation in some seniors with postural instability. Front Neurol. 2016;7(JAN). doi:10.3389/fneur.2016.00004

13. Piker EG, Jacobson GP, Romero D, Wang Y, Smith K. The Clinical Significance of the Failure to Perceive Vertigo in the Postcaloric Period Despite a Robust Caloric Response. Am J Audiol. 2020;29(1):50–58. doi:10.1044/2019_AJA-19-00036

14. Himi T, Kataura A. Vestibular Agnosia in a Patient with Adrenoleukodystrophy. Equilib Res. 1994;53(3):368–373. doi:10.3757/JSER.53.368

15. Jacobson GP, Piker EG, Grantham SL, English LN. Age predicts the absence of caloric-induced vertigo. J Otol. 2018;13(1):5. doi:10.1016/J.JOTO.2017.10.005

16. Harrell R, Manetta C, Guthrie M, Enam N. The Prevalence of Symptom Reporting for Benign Paroxysmal Positional Vertigo in a Traumatic Brain Injury Population. Otol Neurotol. 2023;44(2):172–176. doi:10.1097/MAO.0000000000003770

17. Piker EG, Jacobson GP. Self-report symptoms differ between younger and older dizzy patients. Otol Neurotol. 2014;35(5):873–879. doi:10.1097/MAO.0000000000000391

18. Li Y, Smith RM, Whitney SL, Seemungal BM, Ellmers TJ. We should be screening for benign paroxysmal positional vertigo (BPPV) in all older adults at risk of falling: a commentary on the World Falls Guidelines. Age Ageing. 2023;52(11):1–3. doi:10.1093/AGEING/AFAD206

19. Motin M, Keren O, Groswasser Z, Gordon CR. Benign paroxysmal positional vertigo as the cause of dizziness in patients after severe traumatic brain injury: diagnosis and treatment. Brain Inj. 2005;19(9):693–697. doi:10.1080/02699050400013600

20. Chamelian L, Feinstein A. Outcome after mild to moderate traumatic brain injury: The role of dizziness. Arch Phys Med Rehabil. 2004;85(10):1662–1666. doi:10.1016/j.apmr.2004.02.012

21. Elser H, Gottesman RF, Walter AE, et al. Head Injury and Long-term Mortality Risk in Community-Dwelling Adults. JAMA Neurol. 2023;80(3):260–269. doi:10.1001/JAMANEUROL.2022.5024

22. Hadi Z, Mahmud M, Pondeca Y, et al. The human brain networks mediating the vestibular sensation of self-motion. J Neurol Sci. 2022;443. doi:10.1016/J.JNS.2022.120458

23. Rust HM, Smith RM, Mahmud M, Golding JF, Seemungal BM. Force dependency of benign paroxysmal positional vertigo in acute traumatic brain injury: a prospective study. J Neurol Neurosurg Psychiatry. 2022;93(11):1232–1234. doi:10.1136/JNNP-2022-328997

24. Mayne R. A Systems Concept of the Vestibular Organs. In: Kornhuber HH, ed. Vestibular System Part 2: Psychophysics, Applied Aspects and General Interpretations. Handbook of Sensory Physiology. Springer, Berlin, Heidelberg; 1974:493-580. doi:10.1007/978-3-642-65920-1_14

25. Love J, Selker R, Marsman M, et al. JASP: Graphical Statistical Software for Common Statistical Designs. J Stat Softw. 2019;88(1):1–17. doi:10.18637/JSS.V088.I02

26. Smith SM, Jenkinson M, Johansen-Berg H, et al. Tract-based spatial statistics: voxelwise analysis of multi-subject diffusion data. Neuroimage. 2006;31(4):1487–1505. doi:10.1016/J.NEUROIMAGE.2006.02.024

27. Jenkinson M, Beckmann CF, Behrens TEJ, Woolrich MW, Smith SM. FSL. Neuroimage. 2012;62(2):782-790. doi:10.1016/J.NEUROIMAGE.2011.09.015

28. Isensee F, Schell M, Pflueger I, et al. Automated brain extraction of multisequence MRI using artificial neural networks. Hum Brain Mapp. 2019;40(17):4952. doi:10.1002/HBM.24750

29. Zhang H, Avants BB, Yushkevich PA, et al. High-dimensional spatial normalization of diffusion tensor images improves the detection of white matter differences: an example study using amyotrophic lateral sclerosis. IEEE Trans Med Imaging. 2007;26(11):1585–1597. doi:10.1109/TMI.2007.906784

30. Smith SM, Nichols TE. Threshold-free cluster enhancement: addressing problems of smoothing, threshold dependence and localisation in cluster inference. Neuroimage. 2009;44(1):83–98. doi:10.1016/J.NEUROIMAGE.2008.03.061

31. Gaser C, Dahnke R, Thompson PM, Kurth F, Luders E, Initiative ADN. CAT – A Computational Anatomy Toolbox for the Analysis of Structural MRI Data. bioRxiv. Published online June 13, 2022:2022.06.11.495736. doi:10.1101/2022.06.11.495736

32. Ashburner J, Friston KJ. Voxel-based morphometry--the methods. Neuroimage. 2000;11(6 Pt 1):805-821. doi:10.1006/NIMG.2000.0582

33. Manjón J V., Coupé P, Martí-Bonmatí L, Collins DL, Robles M. Adaptive non-local means denoising of MR images with spatially varying noise levels. J Magn Reson Imaging. 2010;31(1):192–203. doi:10.1002/JMRI.22003

34. Rajapakse JC, Giedd JN, Rapoport JL. Statistical approach to segmentation of single-channel cerebral mr images. IEEE Trans Med Imaging. 1997;16(2):176–186. doi:10.1109/42.563663

35. Tohka J, Zijdenbos A, Evans A. Fast and robust parameter estimation for statistical partial volume models in brain MRI. Neuroimage. 2004;23(1):84–97. doi:10.1016/j.neuroimage.2004.05.007

36. Xia M, Wang J, He Y. BrainNet Viewer: A Network Visualization Tool for Human Brain Connectomics. PLoS One. 2013;8(7):e68910. doi:10.1371/JOURNAL.PONE.0068910

37. Malec JF, Brown AW, Leibson CL, et al. The mayo classification system for traumatic brain injury severity. J Neurotrauma. 2007;24(9):1417–1424. doi:10.1089/NEU.2006.0245

38. Nigmatullina Y, Hellyer PJ, Nachev P, Sharp DJ, Seemungal BM. The neuroanatomical correlates of training-related perceptuo-reflex uncoupling in dancers. Cereb Cortex. 2015;25(2):554–562. doi:10.1093/cercor/bht266

39. Kaski D, Quadir S, Nigmatullina Y, Malhotra PA, Bronstein AM, Seemungal BM. Temporoparietal encoding of space and time during vestibular-guided orientation. Brain. 2016;139(2):392–403. doi:10.1093/brain/awv370

40. Seemungal BM. The cognitive neurology of the vestibular system. Curr Opin Neurol. 2014;27(1):125–132. doi:10.1097/WCO.0000000000000060

41. Seemungal BM, Rizzo V, Gresty MA, Rothwell JC, Bronstein AM. Posterior parietal rTMS disrupts human Path Integration during a vestibular navigation task. Neurosci Lett. 2008;437(2):88–92. doi:10.1016/j.neulet.2008.03.067

42. Seemungal BM. The Mechanisms and Loci of Human Vestibular Perception. University of London; 2006. Accessed December 8, 2020. https://discovery.ucl.ac.uk/id/eprint/1445054/

43. Wirth AM, Frank SM, Greenlee MW, Beer AL. White Matter Connectivity of the Visual-Vestibular Cortex Examined by Diffusion-Weighted Imaging. Brain Connect. 2018;8(4):235–244. doi:10.1089/brain.2017.0544

44. Kirsch V, Keeser D, Hergenroeder T, et al. Structural and functional connectivity mapping of the vestibular circuitry from human brainstem to cortex. Brain Struct Funct. 2016;221(3):1291–1308. doi:10.1007/s00429-014-0971-x

45. Taubert M, Draganski B, Anwander A, et al. Dynamic Properties of Human Brain Structure: Learning-Related Changes in Cortical Areas and Associated Fiber Connections. J Neurosci. 2010;30(35):11670. doi:10.1523/JNEUROSCI.2567-10.2010

46. Welniarz Q, Dusart I, Roze E. The corticospinal tract: Evolution, development, and human disorders. Dev Neurobiol. 2017;77(7):810–829. doi:10.1002/DNEU.22455

47. Ji G, Ren C, Li Y, et al. Regional and network properties of white matter function in Parkinson’s disease. Hum Brain Mapp. 2019;40(4):1253–1263. doi:10.1002/hbm.24444

48. Cousins S, Kaski D, Cutfield N, et al. Vestibular Perception following Acute Unilateral Vestibular Lesions. PLoS One. 2013;8(5):e61862. doi:10.1371/JOURNAL.PONE.0061862

49. De Simoni S, Grover PJ, Jenkins PO, et al. Disconnection between the default mode network and medial temporal lobes in post-traumatic amnesia. Brain. 2016;139(12):3137. doi:10.1093/BRAIN/AWW241

50. Mori S, Oishi K, Jiang H, et al. Stereotaxic white matter atlas based on diffusion tensor imaging in an ICBM template. Neuroimage. 2008;40(2):570–582. doi:10.1016/j.neuroimage.2007.12.035

51. Oishi K, Zilles K, Amunts K, et al. Human brain white matter atlas: Identification and assignment of common anatomical structures in superficial white matter. Neuroimage. 2008;43(3):447–457. doi:10.1016/j.neuroimage.2008.07.009

52. Hua K, Zhang J, Wakana S, et al. Tract Probability Maps in Stereotaxic Spaces: Analyses of White Matter Anatomy and Tract-Specific Quantification. Neuroimage. 2008;39(1):336. doi:10.1016/J.NEUROIMAGE.2007.07.053

53. Wakana S, Caprihan A, Panzenboeck MM, et al. Reproducibility of quantitative tractography methods applied to cerebral white matter. Neuroimage. 2007;36(3):630–644. doi:10.1016/J.NEUROIMAGE.2007.02.049

