## Supplementary file for "Vestibular agnosia is linked to worse balance recovery in traumatic brain injury: a longitudinal behavioural and neuro-imaging study"

### Supplementary material

1) Supplementary methods:
   i. Assessment of peripheral and reflex vestibular function
   ii. Demographics for resting state fMRI
   iii. Resting state fMRI analysis
2) Supplementary results
   i. Functional connectivity link to vestibular recovery
   ii. Non-linear influence of VA upon dizziness (DHI) – exploratory analysis

#### Supplementary methods:

**Supplementary methods: Assessment of peripheral and reflex vestibular function**

**Table S1. Peripheral and reflex vestibular function of patients**

| **Patient** | **vHIT** | | | **Caloric** | |
| --- | --- | --- | --- | --- | --- |
|  | **Asymmetry**  **(%)** | **L gain** | **R gain** | **RC SPV** | **LC SPV** |
| 1 | 4 | 1.29 | 1.21 | - | - |
| 2 | 6 | 0.96 | 1.09 | - | - |
| 3 | - | - | - | 64°/s | -80°/s |
| 4 | 4 | 0.84 | 0.91 | - | - |
| 5^a^ | - | - | - | - | - |
| 6 | 3 | 1.02 | 1.09 | - | - |
| 7 | 4 | 0.89 | 0.93 | - | - |
| 8 | 2 | 0.91 | 0.93 | - | - |
| 9 | 1 | 0.97 | 0.98 | - | - |
| 10 | 4 | 1.00 | 0.96 | - | - |
| 11 | 2 | 0.78 | 0.81 | - | - |
| 12 | 13 | 0.82 | 0.71 | - | - |
| 13 | 3 | 1.15 | 1.18 | - | - |
| 14 | - | - | - | 47°/s | -38°/s |
| 15 | 1 | 1.11 | 1.09 | - | - |
| 16 | 3 | 1.27 | 1.34 | - | - |
| 17 | 2 | 1.23 | 1.28 | - | - |
| 18 | 2 | 0.88 | 0.92 | - | - |
| 19^b^ | - | - | - | - | - |
| 20 | 5 | 0.98 | 1.09 | - | - |
| 21 | 6 | 1.10 | 0.90 | - | - |
| 22 | 6 | 0.89 | 1.00 | - | - |
| 23 | 3 | 1.14 | 1.07 | - | - |
| 24 | 3 | 0.97 | 1.03 | - | - |
| 25 | 6 | 1.11 | 0.98 | - | - |
| 26^c^ | - | - | - | - | - |
| 27 | 7 | 1.44 | 1.25 | - | - |
| 28 | 4 | 0.96 | 1.04 | - | - |
| 29 | 10 | 1.32 | 1.08 | - | - |
| 30 | 7 | 0.93 | 1.06 | - | - |
| 31 | 7 | 0.84 | 0.96 | - | - |
| 32 | 9 | 1.12 | 0.93 | - | - |
| 33 | 2 | 1.05 | 1.10 | - | - |
| 34 | 6 | 0.98 | 1.10 | - | - |
| 35 | 2 | 1.07 | 1.12 | - | - |
| 36 | 3 | 1.31 | 1.23 | - | - |
| 37^d^ | - | - | - | - | - |
| 38 | 1 | 0.90 | 0.92 | - | - |
| 39 | 3 | 1.04 | 1.10 | - | - |

vHIT: video head impulse test; L: left; R: right; RC: left canal; LC: right canal; SPV: slow phase velocity; -: dash indicates that the test was not completed or adequate recording not obtained.

^a^90°/s rotation gains: 0.71 R, 0.58 L; 90°/s stop gains: 0.51 R, 0.50 L

^b^90°/s rotation gains: 0.66 R, 0.67 L; 90°/s stop gains: 0.65 R, 0.67 L

^c^90°/s rotation gains: 0.71 R, 0.85 L; 90°/s stop gains: 0.83 R, 0.74 L

^d^90°/s rotation gains: 1.01 R, 0.89 L; 90°/s stop gains: 0.76 R, 0.76 L

**Supplementary methods: Demographics for resting state fMRI**

As described in the main manuscript, resting-state fMRI analysis from N = 17 (of N = 27) patients was performed as N = 11 patients were removed either due to mismatched field of view parameters or outlier scans. The demographics of these subset of patients (N = 17) are also provided here (Table S2) to facilitate interpretation. Since all 17 of these patients are part of the complete sample of N = 34, we report the same subject number of patients as reported in Table 1 of the main manuscript. Notably in this subsample, 6 of 17 patients had vestibular agnosia. Average age of this subsample was 38.23 ± 13.07 (3 Females), which is close to the complete sample. Table S1 also contains the data of these patients’ peripheral vestibular assessment.

**Table S2.** **Demographics of sub-group of patients used in resting-state fMRI analysis**

| Subject | MOI | Severity MAYO | PTA | VA |
| --- | --- | --- | --- | --- |
| **12** | RTA | Mod-Sev | 1 | 1 |
| **17** | Fall | Mod-Sev | 0 | 0 |
| **18** | RTA | Mod-Sev | 0 | 1 |
| **19** | Fall | Mod-Sev | 0 | 0 |
| **21** | Fall | Mod-Sev | 0 | 0 |
| **23** | Fall | Mod-Sev | 1 | 1 |
| **25** | Fall | Mod-Sev | 1 | 0 |
| **27** | Assault | Mod-Sev | 1 | 0 |
| **29** | Fall | Mod-Sev | 0 | 0 |
| **31** | Fall | Mod-Sev | 0 | 0 |
| **32** | RTA | Mod-Sev | 1 | 1 |
| **33** | Assault | Mod-Sev | 1 | 1 |
| **35** | Fall | Mild-Prob | 0 | 0 |
| **36** | RTA | Mod-Sev | 1 | 1 |
| **37** | RTA | Mod-Sev | 1 | 0 |
| **38** | Fall | Mod-Sev | 1 | 0 |
| **39** | RTA | Mod-Sev | 1 | 0 |

1/0: present/absent; MOI: mode of injury; Mod-Sev: moderate to severe; RTA: road traffic accident

**Supplementary methods: Resting state fMRI analysis**

#### Image acquisition:

Structural and functional MRI images were acquired at time 0 and 6 months using a 3 T Siemens Verio MRI scanner, using a 32-channel head coil. The scanning protocol included: (i) 3D T1-weighted images acquired using MPRAGE sequence (image matrix: 256 × 256; voxel size: 1 × 1; Slices: 160; field of view: 256 × 256 mm; slice thickness: 1 mm; TR = 2300 ms; TE: 2.98 ms); (ii) T2*-weighted images sensitive to blood oxygenation level dependent (BOLD) signal for resting state fMRI (image matrix: 64 × 64; voxel size: 3 × 3 × 3 mm^3^; Slices: 35; field of view: 192 × 192 mm; flip angle: 80°; slice thickness: 3 mm; TR = 2000 ms; TE: 30 ms; volumes = 300; scan time = 10 min); (iii) DTI sequences were acquired in 64 directions with an isotropic voxel size of 2mm^3^, b = 1000 s/mm^2^ with four images with b = 0 s/mm^2^ ( field of view = 256 × 256 mm; matrix size = 128 × 128; TR = 9500 ms; TE = 103 ms). Subjects were instructed to keep their eyes closed, stay awake, and avoid dwelling on any thoughts during resting-state imaging.

#### Functional MRI assessing whole brain connectivity - Resting state (rsfMRI)

##### Preprocessing

Data were pre-processed using the CONN Toolbox^1^ based on Statistical Parametric Mapping software (SPM12; http://www.fil.ion.ucl.ac.uk/spm/). Pre-processing steps were as follows: (1) Realignment to mean functional image, unwarping, and susceptibility distortion correction. (2) Slice timing correction. (3) Functional outlier detection using ART (artifact detection toolbox), ^1^ with scans exceeding a framewise displacement threshold of 0.9 mm were labelled as outliers. (4) Structural segmentation and normalization: Indirect functional normalization using deformation fields estimated from structural normalization. Indirect functional normalization did not work for some participants; for those participants, both functional and structural scans were directly normalized and transformed into MNI space. (5) The individuals' segmented masks were binarized with grey-matter: p > 0.2, white-matter: p > 0.5 (with mask eroded by 1 voxel), and cerebrospinal fluid (CSF): p > 0.5 (with mask eroded by 1 voxel). (6) Smoothing with FWHM of 8 mm. (7) Subsequently, denoising was performed in which six motion regressors (3 translational and 3 angular motion), their temporal derivatives, mean CSF, mean white-matter signal, and outlier scans identified by ART toolbox were regressed out. (8) The residual (denoised) signal was also linearly detrended and band-pass filtered in a frequency range of 0.008–0.1 Hz. Global signal was not regressed out since the global signal regression is known to introduce negative correlations.^2^ (9) Analysis space mask: multiple participants had missing parts of cerebellum due to field of view limitations; thus, cerebellum was removed from the group analysis mask using cerebellum ROI from the CONN software atlas in the SPM software (“imcalc” tool).

##### Independent component analysis

Group independent component analysis (ICA) was performed to assess the intra-network resting state changes associated with the recovery of behavioural measures using Fast ICA algorithm in CONN toolbox. We previously found that estimation of optimal number of independent components (ICs) via algorithms, such as modified minimum description length algorithm, can result in quite high number of ICs.^3^ Thus, similar to our previous report,^3^ we estimated 15 ICs for group level analysis to reduce overfitting and sub-division of ICs, and to avoid statistical evaluation of multiple ICs.

##### Group level analysis

Change in the vestibular perceptual thresholds (VPTs) from acute visit to 6-month visit was estimated and converted into z-score. Similarly for postural balance measure, change in root mean square (RMS) sway in the “soft surface-eyes closed” balance condition from acute visit to 6-month visit was estimated and converted into z-score. These normalized change values of both behavioural measures, vestibular perceptual thresholds and the RMS sway in “soft surface-eyes closed” condition, were used as covariates to identify the interaction of recovery of vestibular perception and postural balance as well as their respective main effects using CONN toolbox.

**Statistical Analysis**

All group level changes in ICA were evaluated using parametric statistics. Findings are reported after correcting for multiple comparisons at cluster-level using both, FDR and FWE correction at *P* < 0.05. Findings are also reported after correction for the statistical evaluation of 15 ICs (corrected *P* = 0.0033). Cluster height threshold was selected as *P* < 0.001.^4,5^ The findings are reported as MNI coordinates (MNI: x, y, z).

#### Supplementary Results:

i. **Functional connectivity link to vestibular recovery (group Independent Component Analysis)**

We performed group independent component analysis (ICA) to identify within-individual functional MRI resting-state networks (RSNs) to assess the link between the change (Δ) in connectivity over time and Δ in VPTs (vestibular perceptual thresholds) and Δ sway and found a significant interaction (FWE corrected *P* < 0.05) as shown in Fig S1 and Table S3. This significant interaction was found in one cluster within IC - 4 (composed of bilateral superior and middle temporal and frontal gyri). The cluster was located at left temporal occipital fusiform cortex. However, it did not survive the conservative correction for the total independent components (*P* < 0.0033).

The main effect of Δ sway (details in Table S3) was found to be significant in two independent components (IC 4 – RSN composed of bilateral superior and middle temporal and frontal gyri and IC 11 – a visual RSN). The main effect of Δ vestibular perceptual thresholds (details in Table S3) was found to be significant in six of the independent components (IC 2 – RSN composed of bilateral superior and middle temporal and frontal gyri; IC 7 – medial visual RSN; IC 8 – Frontoparietal RSN; and IC 11 – visual RSN; IC 14 – Sensorimotor RSN; IC 15 – Subcortical & White-matter RSN).

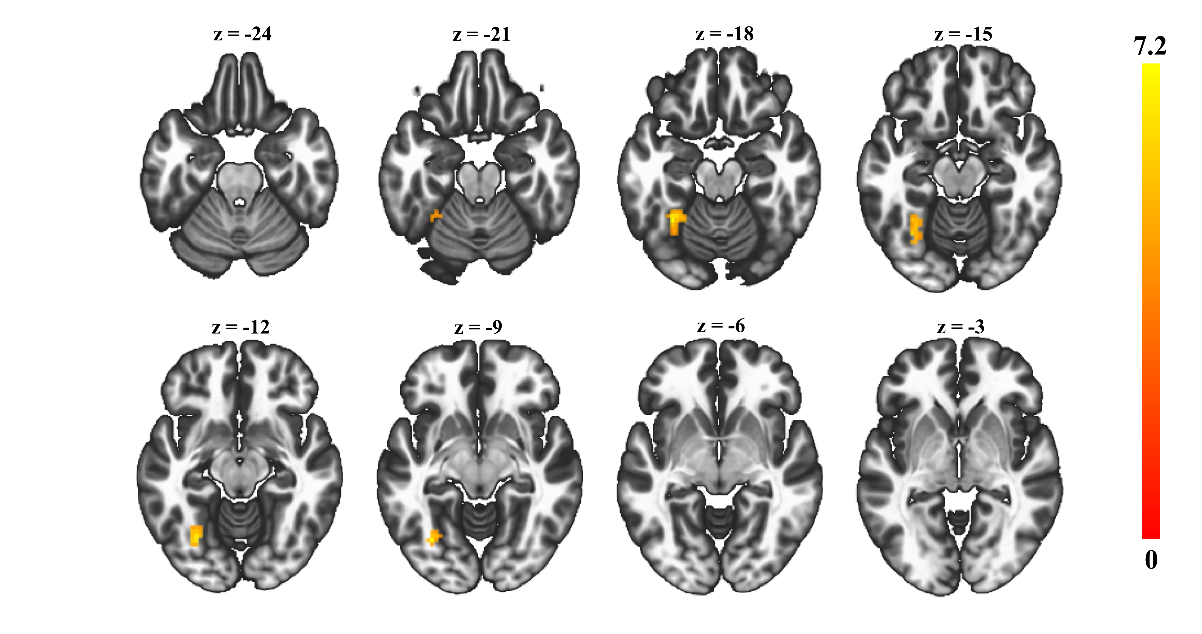

**Figure S1.** **Resting state fMRI results.** The significant interaction (Δ VPTs × Δ sway × Δ functional connectivity) was localised at left temporo-occipital fusiform cortex. (Colour-bar indicate t-values; voxel level P < 0.001 & cluster level FWE corrected P < 0.05). VPTs: vestibular perceptual thresholds; Δ: change.

**Table S3. Resting state imaging (independent component analysis)**

| **Interaction of Δ VPT, Δ sway, and Δ functional connectivity values over time** | | | | |
| --- | --- | --- | --- | --- |
| 1C-4 | Left temporal occipital fusiform cortex | 62 | -33, -51, -18 | t = 7.44^*^ |
| **Conditional main effect of Δ sway** | | | | |
| 1C-4 (1)^c^ | Right precentral gyrus | 52 | 60, -03, 18 | t = -7.24^*^ |
| 1C-11 (1)^c^ | Right frontal pole | 92 | 48, 39, 30 | t = 7.69^***^ |
| 1C-11 (2)^c^ | Cingulate gyrus (posterior) | 80 | 00, -45, 36 | t = -7.14^***^ |
| **Conditional main effect of Δ VPT** | | | | |
| IC-2 | Brain stem (MCP, PCT, CST) | 276 | -09, -30, -33 | t = 9.01^***^ |
| IC-7 | Left frontal pole | 55 | -27, 48, 03 | t = -5.77^*^ |
| IC-8 | Left frontal pole | 53 | -12, 54, 30 | t = -7.29^*^ |
| IC-11(1^a^) | Precuneous | 249 | 00, -45, 36 | t = 8.33^***^ |
| IC-11(2^a^) | Right inferior frontal gyrus, pars opercularis | 139 | 57, 12, 12 | t = -8.85^***^ |
| IC – 11(3^a^) | Right supramarginal gyrus (posterior) | 138 | 60, -39, 42 | t = -9.47^***^ |
| IC – 11(4^a^) | Left lateral occipital cortex (superior) | 67 | -51, -69, 39 | t = 5.70^***^ |
| IC – 11(5^a^) | Right supplementary motor cortex | 47 | 09, 06, 57 | t = -5.30^*^ |
| IC – 11(6^a^) | Left superior frontal gyrus | 43 | -21, 27, 60 | t = 8.22^*^ |
| IC – 11(7^a^) | Precuneus | 38 | -09, -60, 18 | t = 5.36^*^ |
| IC – 14(1^a^) | Right precentral gyrus | 59 | 45, -12, 60 | t = 9.39^*^ |
| IC – 14(2^a^) | Brain stem | 48 | 21, -18, -39 | t = -5.31^*^ |
| IC – 15(1^a^) | Left temporal pole, left hippocampus, left amygdala | 218 | -15, -15, -15 | t = 8^***^ |
| IC – 15(2^a^) | Right precentral gyrus | 215 | 06, -12, 75 | t = 5.81^***^ |
| IC – 15(3^a^) | Right amygdala | 58 | 15, -03, -18 | t = 5.59^*^ |
| IC – 15(4^a^) | Right temporal pole | 48 | 60, 12, -09 | t = 6.66^*^ |

IC: independent component; MCP: middle cerebellar peduncle; PCT: pontine crossing tract; CST: corticospinal tract.

^*^FWE Corrected *P* < 0.05.

^***^FWE corrected *P* < 0.0033 (corrected for 15 independent components analysed).

^a^Independent component (cluster number)

#### Supplementary Discussion:

Using within-patient analyses, we found that the functional brain imaging changes linked to vestibular recovery were anatomically and functionally mirrored to that in acute VA.^3^ Specifically, in the acute phase, ***reduced*** functional connectivity in the right temporal occipital fusiform cortex (right TOFC) correlated with vestibular agnosia (VA),^3^ whereas at follow-up, ***increased*** functional connectivity in left TOFC was linked with vestibular recovery. These findings could imply an interhemispheric interplay between right and left TOFC mediates vestibular recovery of balance and perception following TBI. Given notions of a right hemisphere dominance mediating vestibular function,^6–8^ one speculative interpretation of our data is that left hemispheric TOFC overactivity at follow-up reflects an active compensatory process redressing the acute decrease in right hemispheric TOFC activity.

#### Supplementary Results:

ii. **Non-linear influence of VA upon dizziness (DHI)**

Whilst we did not find any obvious link between dizziness scales and vestibular perceptual thresholds for mild-moderate VA, for severe VA (vestibular perceptual thresholds 3deg/s^2^ or greater) there was a trend showing a non-linear link (Figure S2) when we correlated acute VPTs with acute DHI, providing some weak support for this hypothesis, however this requires additional assessment in future studies. We believe it is worthwhile exploring this non-linear link in future larger studies on the basis that this hypothesis may make useful clinical predictions that can aid clinical care, e.g. increased dizziness over time following TBI may occur with improved brain functioning as perceptual functioning improves.

This non-linear link shown for DHI and VPTs acutely for severe vestibular agnosia was based upon a cluster analysis linking PTA with VPTs (see Figure S3).

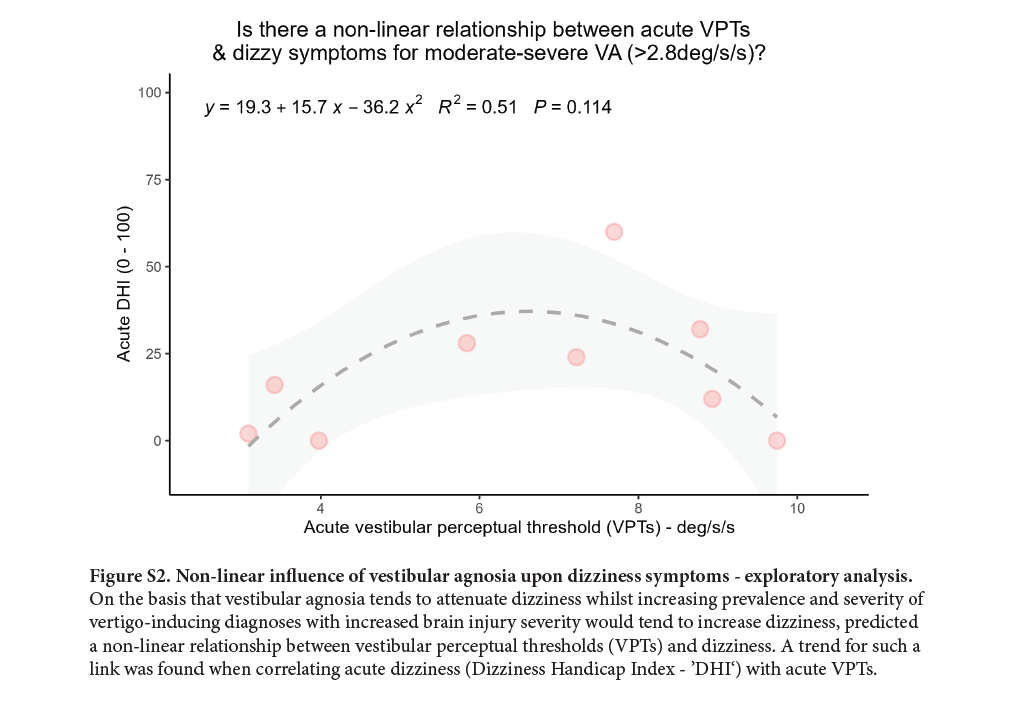

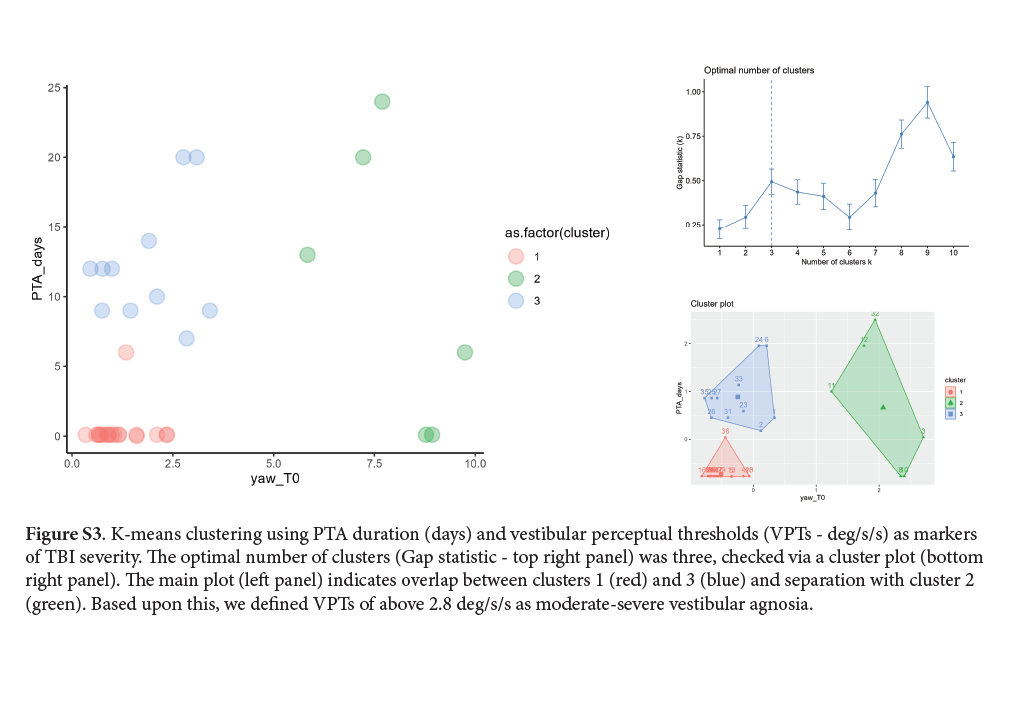

**References**

1.	Whitfield-Gabrieli S, Nieto-Castanon A. Conn: A Functional Connectivity Toolbox for Correlated and Anticorrelated Brain Networks. *Brain Connect*. 2012;2(3):125-141. doi:10.1089/brain.2012.0073

2.	Murphy K, Birn RM, Handwerker DA, Jones TB, Bandettini PA. The impact of global signal regression on resting state correlations: Are anti-correlated networks introduced? *Neuroimage*. 2009;44(3):893-905. doi:10.1016/j.neuroimage.2008.09.036

3.	Hadi Z, Mahmud M, Pondeca Y, et al. The human brain networks mediating the vestibular sensation of self-motion. *J Neurol Sci*. 2022;443. doi:10.1016/J.JNS.2022.120458

4.	Eklund A, Nichols TE, Knutsson H. Cluster failure: Why fMRI inferences for spatial extent have inflated false-positive rates. *Proc Natl Acad Sci U S A*. 2016;113(28):7900-7905. doi:10.1073/PNAS.1602413113

5.	Eklund A, Knutsson H, Nichols TE. Cluster failure revisited: Impact of first level design and physiological noise on cluster false positive rates. *Hum Brain Mapp*. 2019;40(7):2017-2032. doi:10.1002/hbm.24350

6.	Dieterich M, Bucher SF, Seelos KC, Brandt T. Horizontal or vertical optokinetic stimulation activates visual motion-sensitive, ocular motor and vestibular cortex areas with right hemispheric dominance. An fMRI study. *Brain*. 1998;121(8):1479-1495. doi:10.1093/BRAIN/121.8.1479

7.	Dieterich M, Bense S, Lutz S, et al. Dominance for Vestibular Cortical Function in the Non-dominant Hemisphere. *Cereb Cortex*. 2003;13(9):994-1007. doi:10.1093/CERCOR/13.9.994

8.	Dieterich M, Kirsch V, Brandt T. Right-sided dominance of the bilateral vestibular system in the upper brainstem and thalamus. *J Neurol*. 2017;264(1):55-62. doi:10.1007/s00415-017-8453-8
